# COVIDOSE: Low-dose tocilizumab in the treatment of Covid-19

**DOI:** 10.1101/2020.07.20.20157503

**Authors:** Garth W. Strohbehn, Brian L. Heiss, Sherin J. Rouhani, Jonathan A. Trujillo, Jovian Yu, Alec J. Kacew, Emily F. Higgs, Jeffrey C. Bloodworth, Alexandra Cabanov, Rachel C. Wright, Adriana K. Koziol, Alexandra Weiss, Keith Danahey, Theodore G. Karrison, Cuoghi C. Edens, Iazsmin Bauer Ventura, Natasha N. Pettit, Bhakti K. Patel, Jennifer Pisano, Mary E. Strek, Thomas F. Gajewski, Mark J. Ratain, Pankti D. Reid

## Abstract

**Background:** Interleukin-6 (IL-6)-mediated hyperinflammation may contribute to the high mortality of coronavirus disease 2019 (Covid-19). Tocilizumab, an IL-6 receptor blocking monoclonal antibody, has been repurposed for Covid-19, but prospective trials and dose-finding studies in Covid-19 are lacking.

**Methods:** We conducted a phase 2 trial of low-dose tocilizumab in hospitalized adult patients with Covid-19, radiographic pulmonary infiltrate, fever, and C-reactive protein (CRP) ≥ 40 mg/L who did not require mechanical ventilation. Dose cohorts were determined by a trial Operations Committee, stratified by CRP and epidemiologic risk factors. A range of doses from 40 to 200 mg (low-dose tocilizumab) was evaluated, with allowance for one repeat dose at 24-48 hours. The primary objective was to assess the relationship of dose to fever resolution and CRP response. Outcomes were compared with retrospective controls with Covid-19. Correlative studies evaluating host antibody response were performed in parallel.

**Findings:** A total of 32 patients received low-dose tocilizumab. This cohort had improved fever resolution (75·0% vs. 34·2%, p = 0·001) and CRP decline (86·2% vs. 14·3%, p < 0·001) in the 24-48 hours following drug administration, as compared to the retrospective controls (N=41). The probabilities of fever resolution or CRP decline did not appear to be dose-related (p=0·80 and p=0·10, respectively). Within the 28-day follow-up, 5 (15·6%) patients died. For patients who recovered, median time to clinical recovery was 3 days (IQR, 2-5). Clinically presumed and/or cultured bacterial superinfections were reported in 5 (15·6%) patients. Correlative biological studies demonstrated that tocilizumab-treated patients produced anti-SARS-CoV-2 antibodies comparable to controls.

**Interpretation:** Low-dose tocilizumab was associated with rapid improvement in clinical and laboratory measures of hyperinflammation in hospitalized patients with Covid-19. Results of this trial and its correlative biological studies provide rationale for a randomized, controlled trial of low-dose tocilizumab in Covid-19.

**Funding:** ClinicalTrials.gov number NCT04331795. Study infrastructure was supported by NIH CTSA UL1 TR000430.

**Research in Context:** *Evidence before this study:* Many patients with novel coronavirus disease 2019 (Covid-19) develop acute lung injury and hypoxic respiratory failure possibly due to a hyperinflammatory state similar to the cytokine release syndrome that occurs as a complication of chimeric antigen receptor T-cell therapy. Interleukin-6 (IL-6) has been implicated in both processes, leading to the hypothesis that patients with Covid-19 may benefit from IL-6 axis-directed therapies such as the IL-6 receptor-blocking monoclonal antibody tocilizumab. No dose-finding studies have been performed for tocilizumab in the setting of Covid-19.

*Added value of this study:* This prospective phase 2 clinical trial is, to our knowledge, the first to evaluate low-dose tocilizumab in patients with Covid-19 and the first to evaluate the effect of tocilizumab on anti-SARS-CoV-2 antibody response.

*Implications of all the available evidence:* The COVIDOSE study, together with retrospective and real-world evidence studies demonstrating the efficacy of tocilizumab, suggests that low-dose tocilizumab is a potential treatment for hyperinflammation among patients with Covid-19 and merits randomized, controlled testing in this patient population.

## Background

The global pandemic of coronavirus disease-2019 (Covid-19), the disease caused by SARS-CoV-2, threatens public health, with quoted mortality among hospitalized patients exceeding 15%.^1,2^ Covid-19 is hypothesized to progress in three stages. The first stage is marked by localized viral replication producing constitutional symptoms and dry cough, the second by overlapping viral replication and host pulmonary inflammatory response producing dyspnea with or without hypoxia, and finally, a systemic hyperinflammatory stage marked by shock and respiratory failure, associated with high levels of C-reactive protein (CRP) and cytokines, including interleukin-1 (IL-1) and interleukin-6 (IL-6).^3,4^ Repurposed investigational therapies to this point have been divided into antiviral therapies such as lopinavir/ritonavir and remdesivir and also adjunctive anti-inflammatory therapies such as corticosteroids, hydroxychloroquine, Janus kinase inhibitors, Bruton’s tyrosine kinase inhibitors, and immunomodulatory monoclonal antibodies.^2,5,6^

In severely and critically ill patients with Covid-19, IL-6-mediated hyperinflammation resembling cytokine release syndrome (CRS) may drive disease mortality,^7^ suggesting that repurposing of anti-IL-6 axis monoclonal antibodies such as tocilizumab, sarilumab, and siltuximab, or anti-IL-1 therapies such as anakinra, warrant investigation.^3,7,8^ Rapid resolution of clinical and biochemical signs of hyperinflammation has been noted following a single, 400 mg dose of tocilizumab in patients with severe to critical Covid-19,^9^ and one multi-center, retrospective case-control study suggests an approximately 40% reduction in risk of invasive ventilation or Covid-19-related mortality following tocilizumab.^10^ Though peer-reviewed manuscripts are not yet available, an investigator-initiated prospective, multi-institutional, randomized, controlled trial evaluating tocilizumab 8 mg/kg in patients with moderate and severe Covid-19 disease (CORIMUNO-TOCI) is reported to be positive.^11^ Industry-sponsored efforts evaluating tocilizumab 8 mg/kg and sarilumab 400 mg in patients with critical Covid-19 disease are ongoing.^12,13^

However, a tocilizumab dose lower than the labeled CRS dose of 8 mg/kg may be advantageous.^14^ Prior studies suggest that a serum tocilizumab concentration as low as 1 μg/mL – less than 1% of the peak concentration achieved with the 8 mg/kg dose – may be sufficient to blunt greater than 95% of IL-6 receptor (IL-6R) signaling.^15^ In patients with Covid-19, high doses of tocilizumab may increase the risk of secondary bacterial infections (e.g., hospital acquired and ventilator associated pneumonias) approximately three-fold;^10,16^ lower doses may reduce the risk of secondary infection.^14^ Additionally, IL-6 stimulates B-cell proliferation, plasma cell maturation, and antibody responses.^17-19^ Blocking IL-6R with tocilizumab might impair the generation of antibody responses to SARS-CoV-2. Therefore, administration of high-dose tocilizumab too early in the disease has the potential to suppress the adaptive immune response to SARS-CoV-2. Finally, using lower doses of tocilizumab may extend available supplies.^20^

To our knowledge, no dose-finding studies have been conducted for tocilizumab in Covid-19. Drawing on lessons from interventional pharmacoeconomics,^21,22^ we hypothesized that doses of tocilizumab lower than those used in the outpatient rheumatologic (4-8 mg/kg) or the Covid-19 settings (400 mg or 8 mg/kg) would be effective in reducing Covid-19-related inflammation.^14^ We developed a titration regimen to administer low-dose tocilizumab to hospitalized patients with Covid-19.^14^ We present the results of our phase 2 study, demonstrating pharmacodynamic and clinical evidence of low-dose tocilizumab activity in hospitalized patients with Covid-19 associated hyperinflammation not requiring mechanical ventilation.

## Methods

### Trial Design and Oversight

COVIDOSE was an investigator-initiated, non-randomized, open-label trial performed at a single site (University of Chicago Medicine [UCM]). Details of the low-dose tocilizumab treatment strategy have been described previously.^14^ The University of Chicago sponsored the trial but institutionally had no role in the design, execution, or analysis of this trial. There was no industry involvement in COVIDOSE. Disclosures and conflicts of interest have been reported. COVIDOSE was registered with the National Clinical Trials Registry (NCT04331795).

The UCM institutional review board (IRB) approved the trial protocol. Given the rapidly evolving landscape of Covid-19, the IRB authorized an internal trial operating committee to facilitate dose modifications based on incoming data (full details of this operating structure are available in the COVIDOSE trial protocol and Supplementary Appendix). An internal data monitoring committee oversaw the trial’s progress and adjudicated outcomes, safety signals, and potential protocol deviations in weekly meetings. All authors attest to the validity of the data and adherence to the trial protocol, with reporting of violations where indicated.

### Study Participants

Eligible patients were those hospitalized at UCM, aged 18 years or older, and who had: 1) positive polymerase chain reaction test, most commonly nasopharyngeal, for SARS-CoV-2 RNA, 2) chest radiograph findings such as bilateral ground glass opacities or hazy bilateral infiltrates consistent with viral or atypical pneumonia (as determined by UCM radiologists providing usual clinical care independent of COVIDOSE), 3) documented fever, defined as temperature ≥ 38·0°C in the 24 hours prior to the time of tocilizumab administration as measured by commonly accepted clinical methods (predominantly oral or axillary), and 4) C-reactive protein (CRP) ≥ 40 mg/L. Key exclusion criteria included invasive mechanical ventilation, vasopressor medications, active therapy with biological immunosuppressive or Janus kinase inhibitor medications, and previous receipt of an investigational antiviral agent or off-protocol anti-IL6R therapy. Full eligibility criteria are available in the trial protocol. Patients or their legally authorized representatives provided written or electronic informed consent.

### Trial Procedures

Enrolled patients were subdivided into two groups (Group A and Group B) based on laboratory signs of hyperinflammation and the presence or absence of risk factors for Covid-19-related mortality (based on extant univariate regressions at the time of study and as agreed to by multidisciplinary panel of institutional experts). Risk factors, nearly all of which were subsequently validated,^23^ included: any previous intensive care unit admission; previous non-elective intubation; hospitalization for exacerbation of congestive heart failure or chronic obstructive pulmonary disease in the past 12 months; coronary artery disease requiring percutaneous coronary intervention or coronary artery bypass grafting; stroke with residual neurologic deficit; pulmonary hypertension; home supplemental oxygen use; interstitial lung disease; asthma requiring inhaled corticosteroid use; history of pneumonectomy, lobectomy, or radiation therapy to lung; HIV/AIDS; cancer diagnosis (any stage) receiving non-hormonal treatment; immunodeficiency; end-stage renal disease requiring hemodialysis or peritoneal dialysis; body-mass index > 30 kg/m2; and inpatient supplemental oxygen requirement > 6L/min at the time of enrollment. Group A patients had CRP **≥** 75 mg/L and at least one risk factor for mortality. Group B patients had CRP of 40-74 mg/L or lacked risk factors for Covid-19-related mortality.

After group assignment, patients were treated according to the low-dose tocilizumab algorithm, integrated into usual clinical care (Figure S1).^14^ Group A patients received tocilizumab 200 mg (cohort 1A) or 120 mg (cohort 2A). Group B patients received 80 mg (cohort 1B) or 40 mg (cohort 2B). All dosage levels were protocol-based and determined on a cohort basis by the trial operating committee. Vital signs and laboratory studies were monitored per usual clinical care. CRP was evaluated immediately prior to tocilizumab administration (baseline) and approximately 24 hours following tocilizumab administration, except where detailed. Patients were eligible for re-dosing with tocilizumab. In the originally developed re-dosing schema, the decision was guided strictly by CRP response. In the final version of the protocol, however, the re-dosing decision was guided by biochemical and clinical parameters. Patients were re-dosed with tocilizumab if signs of clinical worsening (as defined by increased supplemental oxygen requirement or worsening fever curve determined by maximum temperature) were accompanied by CRP decline of < 25% when compared to baseline at the 24-hour assessment (Figure S1). In the COVIDOSE algorithm, treating physicians maintained the option of administering off-protocol tocilizumab 400 mg as indicated per their clinical judgment. Please see the Supplementary Appendix for further information.

Patients were followed for the duration of their hospitalization and were contacted 28 days after tocilizumab administration. At the 28-day timepoint, survival, clinical status (including admission to a medical or assisted living facility), new or persistent supplemental oxygen requirement, and any notable diagnoses or treatments for secondary infection were documented. Planned enrollment was fifty patients, with the goal of identifying a pharmacodynamically active, low dose of tocilizumab for the treatment of Covid-19. After enrolling the thirty-second patient, remdesivir received Emergency Use Authorization and quickly became institutional standard of care.^24^ To avoid collecting heterogeneous data confounded by remdesivir use, the COVIDOSE trial operating committee elected to close the study to further enrollment; it was not closed due to safety concerns.

### Retrospective Controls

A retrospective control group was assembled using an institutional Covid-19 clinical data warehouse of de-identified medical records by identifying patients who met all COVIDOSE eligibility criteria (Figure S2). An additional antibody control cohort with blood samples available from the institutional Covid-19 biobank was used to compare antibody responses (Figure S3). Full descriptions of the retrospective control populations’ derivations are available in the Supplementary Appendix.

### Outcomes

The primary clinical outcome was resolution of fever in the 24-hour period following tocilizumab, defied as a maximum temperature (T_max24hrs_) < 38·0°C. Originally, the primary biochemical outcomes were rate of and time to CRP normalization, guided by earlier tocilizumab-related work.^9,14^ During the conduct of the study, it became apparent that a patient could be safely discharged before CRP had normalized, making CRP normalization impractical. We therefore report the percentage of patients who achieved biochemical response, defined as a CRP reduction ≥25% from baseline in the 24-48 hours after tocilizumab administration, consistent with a decline determined by CRP half-life.^9,25^

Secondary outcomes included overall survival at 28 days, survival to hospital discharge, rate and duration of non-elective mechanical ventilation, time to mechanical ventilation, rate and duration of vasopressor/inotropic agent utilization, time to vasopressor/inotropic agent utilization, and number of days spent in intensive care unit. One month after enrollment began, duration of supplemental oxygen requirement greater than a patient’s baseline was added. Definitions of outcomes are provided in the protocol. We do not include results in this manuscript for all endpoints.

### *Post-Hoc* Evaluation of Clinical Recovery

Randomized, controlled trials of Covid-19 therapies had not been published at the time of COVIDOSE initiation but were published prior to COVIDOSE data maturity.^6^ To add context to COVIDOSE results, time to clinical recovery was evaluated post-hoc. Using a previously developed seven-point ordinal scale,^26^ disease severity was assessed daily, with the patient’s worst clinical status and consequent ordinal score recorded for a given day.^6,26^ Recovery was defined as the first day on which a patient achieved a clinical status of either “hospitalization without need for supplemental oxygen or ongoing medical care” or “not hospitalized”.^6,26^ Time to recovery was defined as the time (in days) from study enrollment to achievement of recovery.^6^ The detailed clinical annotations needed to categorize patients were not available for the retrospective control population. Two independently operating authors (GWS, PDR) adjudicated ratings.

### SARS-CoV-2 Spike and RBD Enzyme-Linked Immunosorbent Assays

The SARS-CoV-2 Spike and Receptor Binding Domain (RBD) protein expression constructs were obtained from Florian Krammer and Patrick Wilson,^27^ and used to generate recombinant protein for an enzyme-linked immunosorbent assay (ELISA) adapted from established protocols.^27^ Goat anti-human immunoglobulin (Ig)-Horseradish peroxidase (HRP) reactive to heavy and light chains of human IgG, IgM, and IgA was used as the secondary antibody (SouthernBiotech; Birmingham, AL, USA).

Antibody titers were determined by measuring total immunoglobulin (IgG, IgM, IgA) reactive against the SARS-CoV-2 Spike glycoprotein and its receptor-binding domain (RBD). The Spike glycoprotein, a structural protein of SARS-CoV-2, is a crucial component in the attachment and entry of the virus into host cells. ELISA titers against the spike protein have been reported to correlate significantly with virus neutralization.^27^ Specifically, the RBD protein binds the angiotensin-converting enzyme 2 (ACE2) receptor on human cells, and likely serves as a target for neutralizing antibodies that facilitate virus clearance. Full experimental details available in the supplemental methods.

### Statistical Analysis

We assessed differences in the clinical and biochemical response rates of patients treated with low-dose tocilizumab compared with the retrospective control population using a two-sided Fisher’s exact test. We also assessed differences in the clinical and biochemical response rates between different tocilizumab doses using a two-sided Fisher’s exact test, unadjusted for cohort or other risk factors. A p-value of < 0·05 was considered to indicate statistical significance. All analyses were performed using Stata (Stata Corp., College Station, TX, USA). For post hoc analysis, descriptive data summaries are reported as median (interquartile range) or percentages. Log_10_ titers of anti-SARS-CoV-2 antibodies in the tocilizumab-treated and untreated populations were compared using Welch’s t-test in R (R Foundation for Statistical Computing; Vienna, Austria).

### Findings

#### Trial Population

From April 1 through May 13, 2020, 32 patients consented to participate in COVIDOSE. Twelve were assigned to Group A, 8 of whom received 200mg and 4 of whom received 120mg. Twenty were assigned to Group B, 15 of whom received 80 mg and 5 of whom received 40 mg (Figure 1). Median time from hospital admission to study enrollment was 1 day (IQR, 1-2 days). All patients were included in the statistical analysis of fever resolution, and 29 patients were included in the statistical analysis of CRP response. There were 41 patients in the retrospective control population (Figure S2). The COVIDOSE subpopulations and retrospective control population differed significantly in terms of epidemiologic risk factors for Covid-19-related decompensation and hydroxychloroquine use (Table 1). Re-dosing of low-dose tocilizumab is discussed in the Supplementary Materials.

**Figure 1.**
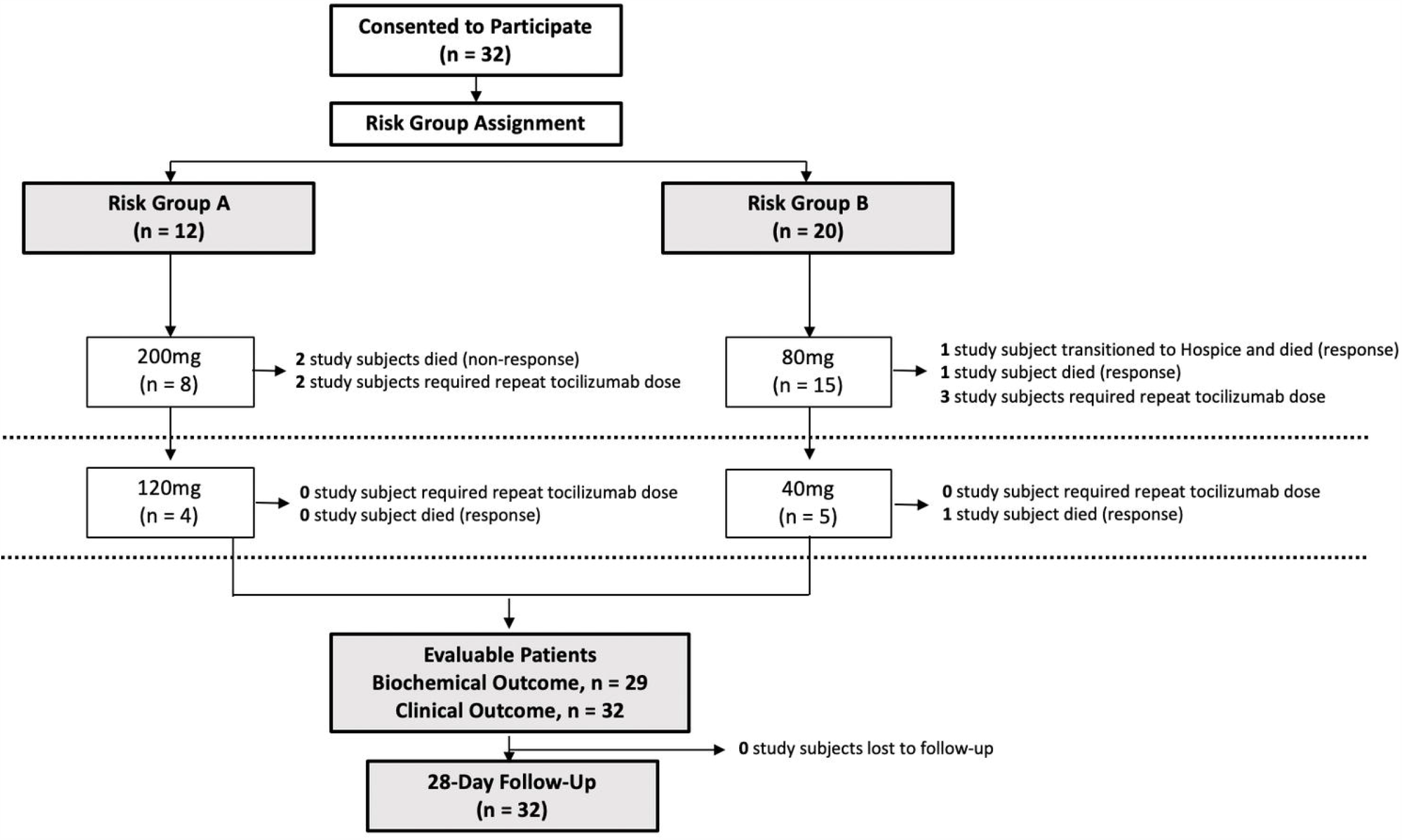
Flow of patients in the COVIDOSE trial. Thirty-two eligible patients consented to participate in the COVIDOSE trial, of which 12 were assigned to Group A and 20 were assigned to Group B on the bases of magnitude of C-reactive protein elevation and epidemiologic risk factors for Covid-19 related mortality. Group A patients received either 200 or 120 mg of tocilizumab and were followed for 28 days following drug administration. Group B patients received either 80 or 40 mg of tocilizumab and were followed for 28 days following drug administration. All 32 patients were evaluable for the purposes of primary clinical outcome and 29 were evaluable for biochemical outcome. No patients were lost to follow-up.

**Table 1.**
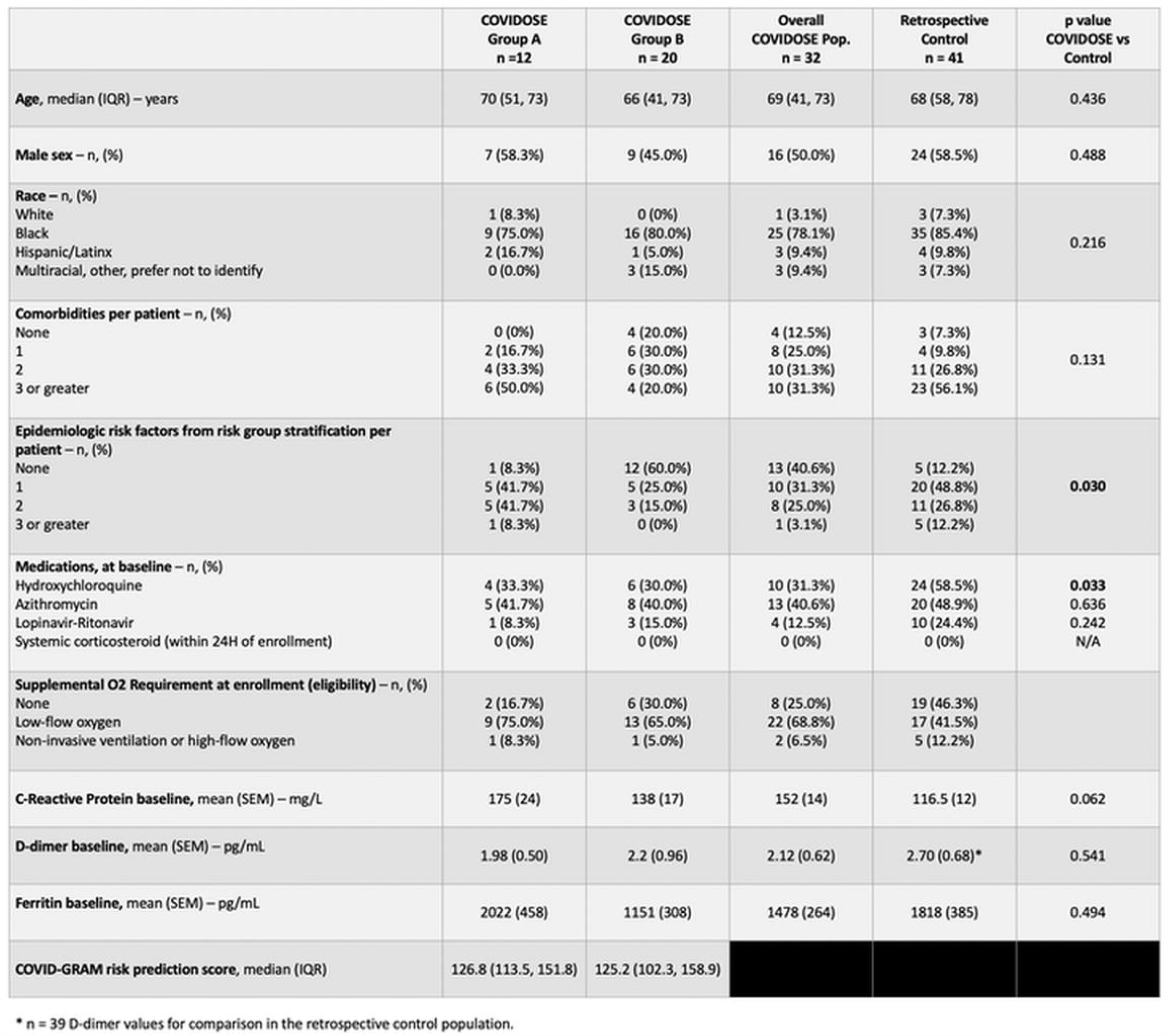
Demographic, clinical, and laboratory characteristics of patients enrolled on COVIDOSE and the retrospective control population. Epidemiologic risk factors include:

#### Clinical Outcomes

Curves of maximum daily temperature (mean ± SE) for the COVIDOSE and retrospective control populations are shown in Figure 2A. At 24 hours following tocilizumab administration, 24 COVIDOSE patients (75·0%) and 14 retrospective control patients (34·1%) achieved T_max24hrs_ < 38·0°C (P = 0·001 for difference compared to control). Eight of 12 (66·7%) Group A patients achieved T_max24hrs_ < 38·0°C (P = 0·055, compared to control) and 16 of 20 (80·0%) Group B patients achieved T_max24hrs_ < 38·0°C (P = 0·001, compared to control) (Figure 2B). Eleven of 15 (73·3%) patients in the 80 mg group achieved T_max24hrs_ < 38·0°C, compared to 5 of 5 (100%) patients in the 40 mg group (P = 0·53) (Figure 2C). T_max24hrs_ < 38·0°C was observed in 5 of 8 (62·5%) patients in the 200 mg group and 3 of 4 (75·0%) patients in the 120 mg group (P = 1·0) (Figure 2C).

**Figure 2.**
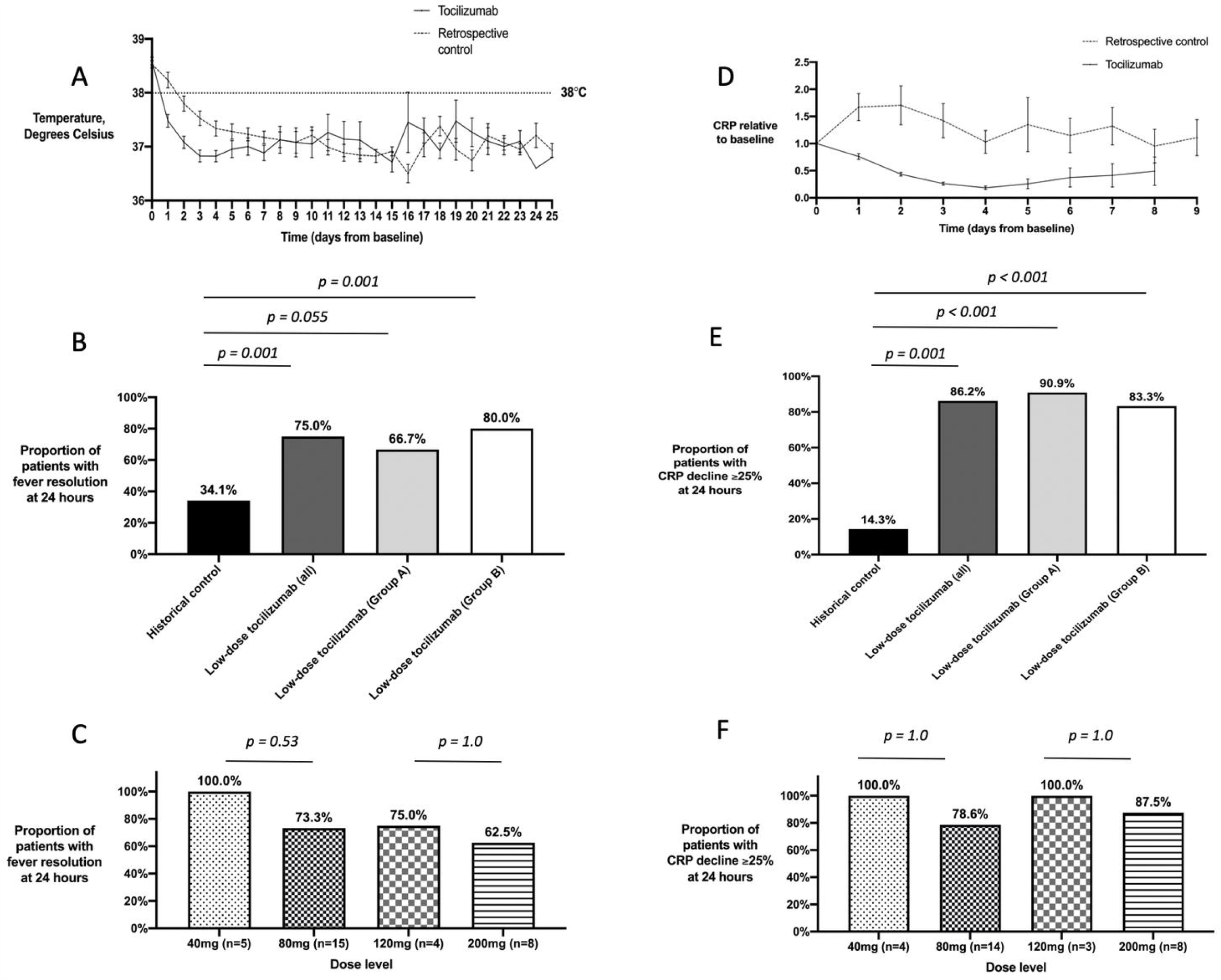
Temperature and C-reactive protein decrease rapidly following administration of low-dose tocilizumab compared to retrospective controls. (A) Temperature values for COVIDOSE patients (closed circle, solid line) and retrospective control patients (open circle, dotted line) following eligibility (mean ± standard error of mean). (B) Percentage of retrospective control (black), COVIDOSE (dark gray), COVIDOSE Group A (light gray), and COVIDOSE Group B (white) patients meeting the primary outcome of T_max24hrs_ < 38·0°C. Bars denote comparisons between groups. (C) Relationship between tocilizumab dose and probability of achieving fever resolution. Percentage of COVIDOSE patients at the noted tocilizumab dose levels achieving the outcome of T_max24hrs_ < 38·0°C. (D) Representative C-reactive protein values for COVIDOSE patients (closed circle, solid line) and retrospective control patients (open circle, dotted line) following eligibility (mean ± standard error of mean). (E) Percentage of retrospective control (black), COVIDOSE (dark gray), COVIDOSE Group A (light gray), and COVIDOSE Group B (white) patients meeting the primary biochemical outcome of CRP decline of ≥ 25% at 24-48 hours. 15 historical control patients with no C-reactive protein readings between 24 and 48 hours were excluded from analysis. Bars denote comparisons between groups. (F) Relationship between tocilizumab dose and probability of achieving CRP decline of ≥ 25%. Percentage of COVIDOSE patients at the noted tocilizumab dose levels achieving CRP decline of ≥ 25%. Bars denote one-sided Fisher’s exact test for difference in proportions. CRP, C-reactive protein. P values are as shown in the figure.

#### Biochemical Outcomes

CRP trends (mean ± SE) for the COVIDOSE and retrospective control populations are shown in Figure 2D. Twenty-eight (68·3%) retrospective control patients had repeat CRP 24-48 hours after qualifying CRP. Twenty-nine (90·6%) COVIDOSE patients had repeat CRP for response assessment 24-48 hours after first low-dose tocilizumab administration. Twenty-five (86·2%) COVIDOSE patients achieved CRP decrease ≥ 25%, compared to 4 (14·3%) retrospective control patients (P < 0·001) (Figure 2E). Ten of 11 (90·9%) Group A patients (P < 0·001) and 15 of 18 (83·3%) Group B patients (P < 0·001) achieved CRP decrease ≥ 25% (Figure 2E). Eleven of 14 (78·6%) patients receiving 80 mg achieved CRP decrease ≥ 25%, compared to 4 of 4 (100%) patients receiving 40 mg (P = 1·0) (Figure 2F). CRP decrease ≥ 25% was observed in 7 of 8 (87·5%) patients receiving 200 mg and 3 of 3 (100%) patients receiving 120 mg (P = 1·0) (Figure 2F).

#### Post-Hoc Evaluation of Recovery Time and Safety

Five of 32 (15·6%) COVIDOSE patients died within 28 days of first tocilizumab administration (Table 2). Median time to recovery for those COVIDOSE patients who recovered was 3 days (IQR, 2-5). Median time to recovery for COVIDOSE patients requiring supplemental oxygen at enrollment (n = 17) was 4 days (IQR, 3-5) (Table 2). Median time to recovery of COVIDOSE patients not requiring supplemental oxygen at enrollment (n = 11) was 2 days (IQR, 2-3) (Table 2). Group A patients’ median time to recovery was 4·5 days (IQR, 2·5-6·75) (Table 2); for Group B patients it was 3 days (IQR, 2-4) (Table 2). Culture-proven or clinically suspected ventilator-associated or hospital-acquired pneumonias were identified in 5 (15·6%) COVIDOSE patients.

**Table 2.** *Post hoc* clinical outcomes in the overall COVIDOSE study population and according to baseline ordinal clinical status and risk group assignment. NIV, non-invasive ventilation; HHFNC, heated high-flow nasal cannula.

|  | Overall | Ordinal Score at Baseline |  |  | Risk Group Stratification at Baseline |  |
| --- | --- | --- | --- | --- | --- | --- |
|  |  | 3 – NIV, HHFNC<br>(n=4) | 4 – Low-flow oxygen<br>(n=17) | 5 – No supp. oxygen<br>(n=11) | Risk Group A<br>(n=12) | Risk Group B<br>(n=20) |
| **Recovery**<br>No. of recoveries<br>Median time to recovery (IQR) – days | 27<br>3 (2-5) | 2<br>11.5 (10.75-12.25) | 15<br>4 (3-5) | 10<br>2 (2-3) | 10<br>4.5 (2.5-6.75) | 17<br>3 (2-4) |
| **Mortality**<br>No. of deaths by day 28 | 5 | 2 | 2 | 1 | 2 | 3 |

#### Tocilizumab does not inhibit the antibody response to SARS-CoV-2

To evaluate whether IL-6 axis blockade impairs the generation of antibody responses to SARS-CoV-2, we analyzed longitudinal blood samples from patients enrolled in a Covid-19 biobanking protocol. Twenty-three patients treated with tocilizumab (n = 19 on COVIDOSE, n = 4 off-protocol 400 mg) and 37 matched control patients were retrospectively identified from the biobank (Figure S3; Table S1). All analyzed patients had pulmonary infiltrate on chest radiograph, temperature ≥ 100·4°F, and CRP ≥ 40 mg/L within 24 hours of tocilizumab administration (Figure S3). Matched control patients were eligible if they met the above criteria (Figure S3). Patients with bacteremia or requiring invasive ventilation were excluded (Figure S3). Patients were not excluded if they had received investigational remdesivir (n = 26; 70·3%).

All patients treated with tocilizumab generated antibodies to the SARS-CoV-2 Spike protein and RBD during infection (Figure 3). Anti-Spike (Figure 3A, 3B) and anti-RBD (Figure 3C, 3D) antibodies from samples 10+ days after symptom onset reached comparable maximum log_10_ titers in tocilizumab-treated and matched control patients (Spike, 4·911 vs 5·000, p = 0·726; RBD, 4·365 vs 4·748, p = 0·149). The rates of titer change from first positive sample to maximum titer within 21 days were not significantly different between tocilizumab-treated and matched control patients (Spike, 1·818 vs 1·298, p = 0·221; RBD, 1·418 vs 1·152, p = 0·412). Although the number of samples in smaller subgroups limits the power of comparisons, there were no differences between COVIDOSE patients and those who received 400 mg (Spike, 4·953 vs 4·745, p = 0·721; RBD, 4·429 vs 4·106, p = 0·48), or between remdesivir-treated or -untreated control patients (Spike, 5·068 vs 4·811, p = 0·63; RBD, 4·867 vs 4·422, p = 0·369) (Figure S4). Taken together, these data suggest that tocilizumab as administered in this study does not impair the induction of anti-SARS-CoV-2 antibodies during infection.

**Figure 3.**
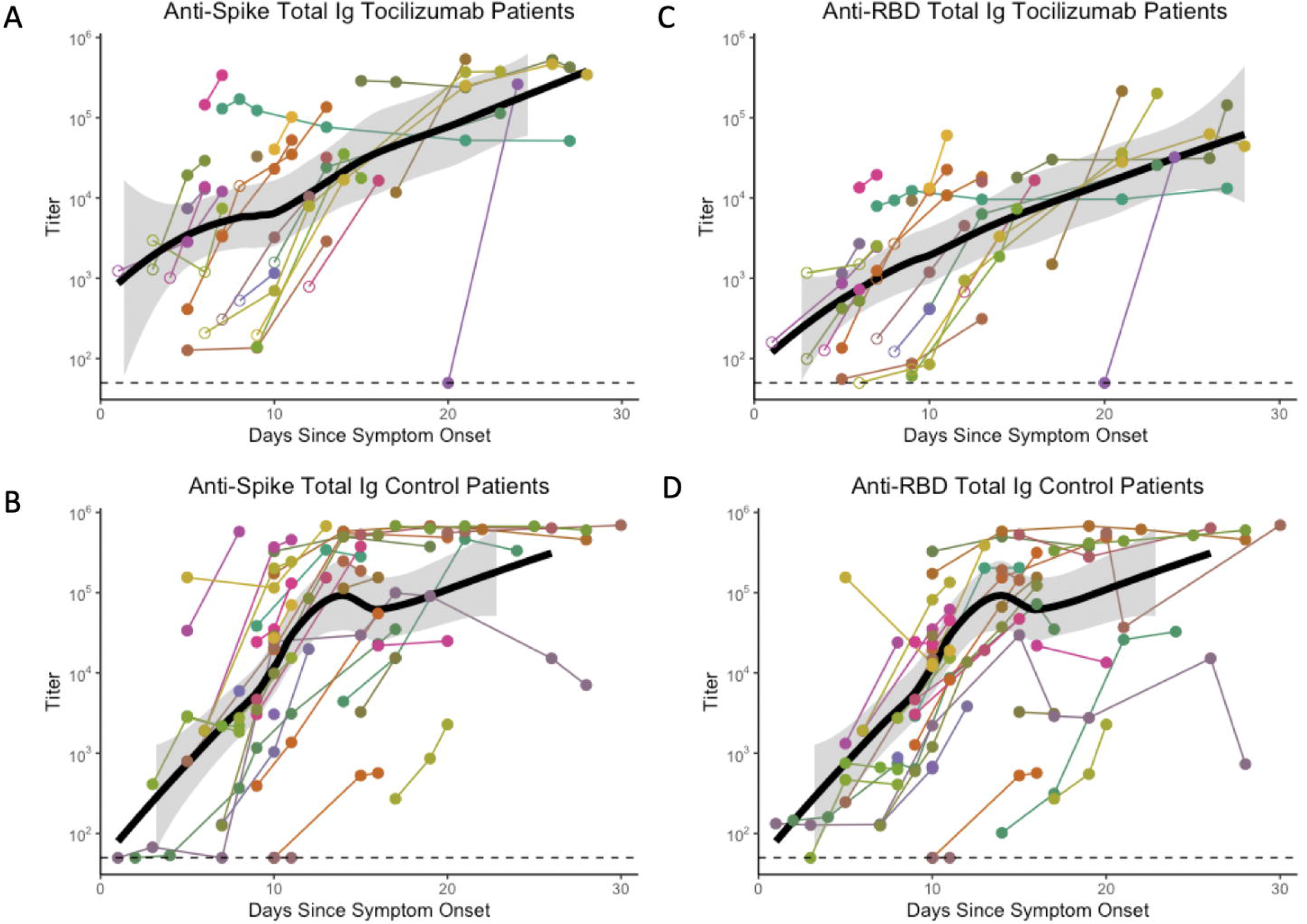
Tocilizumab treated patients develop similar peak antibody levels as control patients. Spike (A,B) and RBD (C,D) antibody titers in tocilizumab-treated (A,C) vs control (B,D) patients over time. Open circles represent time points prior to tocilizumab treatment, and filled circles are after tocilizumab treatment. Each line/color represents a separate patient. Dotted horizontal line represents level of detection in normal donor controls.

### Interpretation

Low-dose tocilizumab was associated with rapid clinical and biochemical improvement in patients with Covid-19 and related hyperinflammation who did not require invasive ventilation, with no apparent relationship between tocilizumab dose and clinical or biochemical improvement. Moreover, there was no apparent diminution in the development of anti-SARS-CoV-2 antibodies in patients who received tocilizumab. Although the minimum effective dose of tocilizumab was not identified, the COVIDOSE study demonstrated that a tocilizumab dose of 40 mg may be sufficient to blunt the clinical and biochemical signs of Covid-19-related hyperinflammation. Referencing clinical pharmacology arguments,^14^ the absence of a dose-response relationship suggests that a dose lower than those utilized in other Covid-19 studies (400 mg) or approved by regulatory bodies for the treatment of CAR-T-related CRS (4-8 mg/kg) may be sufficient. The COVIDOSE trial provides proof-of-concept for a Covid-19 therapeutic strategy centered on the administration of low-dose tocilizumab, with re-dosing guided by readily assessable clinical and biochemical responses.

To our knowledge, COVIDOSE is the first trial to provide prospectively collected evidence of low-dose tocilizumab’s clinical benefit. Marked declines in maximum body temperature and CRP were noted in the 24-48 hours following administration of low-dose tocilizumab – responses that mimic those observed in patients with severe and critical Covid-19 who received tocilizumab 400 mg.^9^ Post hoc analysis of COVIDOSE patients’ clinical trajectories provides encouraging evidence of low-dose tocilizumab’s benefit: Patients requiring supplemental oxygen at enrollment experienced median time to recovery of 4 days, three days shorter than the median time to recovery of a similarly risked subpopulation receiving remdesivir.^6^ The COVIDOSE data will add context to the interpretations of the COVACTA, CORIMUNO-TOCI, and RECOVERY tocilizumab trials when published, as well as the emerging real-world evidence for tocilizumab’s benefit.^10-12,16^

The data presented in COVIDOSE suggest that a three- to ten-fold increase in the effective supply of tocilizumab may be possible. In this trial, we administered a total of 4,120 mg of tocilizumab, corresponding to 37 doses for 32 patients; administering 400 mg to each patient would have required 12,800 mg.^9,11^ Rapid, global scaling is required for tocilizumab to become a safe, effective, and equitable therapy for hospitalized patients with Covid-19 – an interventional pharmacoeconomics-based approach is one possible strategy for extending supplies.^20^

Beyond sustainability, low-dose tocilizumab may be clinically advantageous over higher dosages. The rate of secondary bacterial infection in COVIDOSE was approximately one-fourth that of patients who received tocilizumab 8 mg/kg.^10,16^ Biological studies conducted in parallel with COVIDOSE did not find statistically significant differences in anti-SARS-CoV-2 antibody production between patients treated with tocilizumab (standard or low dose), remdesivir, or supportive care alone. How the prospectively-collected rates of secondary infections compare between low-dose tocilizumab-, tocilizumab 8mg/kg-, and dexamethasone-treated patients, as well as the combination of dexamethasone and tocilizumab, remains to be seen.^2^ Carefully conducted prospective trials with appropriate biological correlative studies are needed.

The COVIDOSE trial is not without limitations. It was a small, single-center study conducted at a tertiary care center with active research efforts in Covid-19-related supportive care. Our trial lacked a contemporaneous, randomized comparator arm, and the higher rate of epidemiologic risk factors in the retrospective control population may have contributed to its slow recovery in terms of both fever resolution and CRP decline. The absence of a randomized control arm limits formal conclusions about low-dose tocilizumab’s safety and efficacy. Finally, the rapidly changing understanding of Covid-19 and local practice prevented trending CRP to normalization, leading to the need for reporting of a different biochemical outcome than originally expected. The COVIDOSE study, however, provides proof-of-concept as well as valuable lessons that will be incorporated into a randomized, controlled trial evaluating the clinical efficacy of low-dose tocilizumab in Covid-19.

## Data Availability

Available upon request for appropriate hypothesis-driven scientific pursuits.

## Acknowledgements

The authors wish to thank Spring Maleckar for her assistance and contributions in creating the REDCap database and Bethany Martell for her expertise throughout aspects of the COVIDOSE trial’s proceedings. We also thank Athalia Pyzer, MD, Yuanyuan Zha, Blake Flood, Glee Li, Hongyuan Jiao, Apameh Pezeshk, Lara Kozloff, Randy Sweis, and the Human Immunologic Monitoring core facility of the University of Chicago Comprehensive Cancer Center for contributions to the Covid-19 biobanking project.

## Individual Funding

Dr. Heiss is supported by the National Institute of General Medical Sciences (T32 GM007019). Dr. Rouhani is supported by the National Institutes of Health (T32 CA009566). Mr. Kacew receives research support from the Pritzker School of Medicine and from the American Society of Hematology. Ms. Higgs is supported by the National Institutes of Health (F30CA250255).

## Supplementary Appendix

## Supplementary Methods

### Biochemical Outcome Measure Determination

COVIDOSE was designed to interdigitate with the provisioning of standard clinical care. Lab ordering therefore was conducted at the discretion of the primary treating physicians, with recommendation made by COVIDOSE investigators. Consequently, CRP results were not uniformly available exactly 24 hours following tocilizumab administration. Much more commonly, results were available between 16- and 48-hours following tocilizumab administration. Given lack of reliability around short-term CRP responses to tocilizumab in either the Covid-19 or rheumatoid arthritis contexts, we report CRP response using laboratory studies collected in the 24- to 48-hour interval after tocilizumab administration. Retrospective control population CRP data was assessed similarly: CRP response is reported using laboratory studies collected 24-48 hours after the patient would have met eligibility criteria for COVIDOSE (baseline CRP).

CRP tends to decline as a patient recovers from Covid-19. Therefore, the time delay between enrollment, baseline CRP collection, and tocilizumab administration may have given COVIDOSE patients a relative “advantage” in the analysis. To explore the possibility further, we also conducted the retrospective control examination with the addition of 6 hours, assessing biochemical response based on CRP studies collected 30-54 hours after first eligible CRP value. 6 hours represented the maximum time delay between baseline CRP measure and drug administration in the COVIDOSE population. This is the comparative retrospective control data presented.

### Retrospective Control Population Derivation

Beginning from a de-identified repository of 2,868 hospitalized patients who tested positive for SARS-CoV-2 RNA by PCR, 449 patients had at least one documented temperature of 100·4°F (38°C) or greater, 508 patients had at least one CRP of greater than or equal to 40 mg/L, and 627 patients had chest imaging study consistent with Covid-19 (as described in Supplementary Methods). 63 patients met all criteria, timed such that the documented CRP ≥ 40 mg/L occurred within 24 hours of documented temperature of 38°C or greater. Patients meeting COVIDOSE exclusion criteria were excluded from the retrospective control population. 22 additional patients were excluded on the bases of receipt of off-label tocilizumab 400 mg prior to occurrence of their first eligible fever, laboratory abnormalities at the time of all eligible fevers, invasive ventilation at the time of eligible fever, the absence of a baseline CRP at the time of first eligible fever, or ineligible age at time of fever/CRP.

### Supervised Machine Learning of Chest Imaging Interpretation

We developed a supervised machine learning algorithm to identify patients with radiographic evidence of infiltrates on chest radiographs or computed tomography, as documented in independent radiologist reports. An initial training data set of 767 imaging study reports was manually annotated by a single rater (J.Y.) as either positive or negative for Covid-19-related infiltrates. These were selected by using either imaging from patients from our intervention arm or imaging from separate admissions for patients who clinically qualified for this trial. In this training set there were 327 negative images and 440 positive images. These classifications were then tabulated with the narrative radiology reads and used as input to develop the supervised machine learning classifier. The reads were converted features using standard natural language processing (NLP) pre-processing, and a random forest NLP classifier was trained to predict presence of infiltrates using this data, validated using repeated k-fold cross-validation.

Algorithm training/classification were performed in R (R Foundation for Statistical Computing; Vienna, Austria) using the ‘tm’ (Text Mining)^1^ and ‘caret’ (Classification And REgression Training)^2^ packages. The radiology report texts were tokenized and stemmed into atomic components by word. These were then filtered using a standard stopword database provided in ‘tm’ as well as an additional list of stopwords curated to ignore descriptions of anatomic or non-clinical features in the radiology reports (see below).

Notably, prior to processing, the adjective form of “no” was concatenated with the subsequent word in all sentences to provide additional context and to prevent it from being removed as a stopword. The random forest algorithm was performed using 50 trees with repeated k-fold cross-validation over k of 10 with 3 repeats. The predicted sensitivity was 94·9% and specificity was 53·3%, with an overall accuracy of 77·2%. This trained classifier was then used on 4510 images to identify 2577 images with evidence of infiltrates in 660 unique patients.

### Categorizing the Retrospective Control Population

Output included from the time of qualifying fever and all recordings thereafter the following: Pulse oximetry, fraction of inspired oxygen, temperature, and CRP. Fraction of inspired oxygen nearest to the patient’s qualifying fever was used to identify that patient’s oxygenation status as either none, low-flow oxygen, or non-invasive ventilation or high-flow oxygen. Receipt of the following medications were identified for each qualifying retrospective control patient in the qualifying hospital encounter: Lopinavir/ritonavir, hydroxychloroquine, and azithromycin, which were alternative Covid-19 treatments at the time of the COVIDOSE trial. Receipt of tocilizumab at a timepoint after the qualifying fever was noted for each patient. Baseline CRP was determined as the first known CRP ≥ 40 mg/L that was collected within 24 hours of the patient’s qualifying fever. Baseline D-dimer and ferritin were the first recorded values from the patient’s hospitalization.

Using the International Classification of Diseases, 10^th^ Revision, Clinical Modification, risk factors corresponding to the diagnoses used for risk-stratification of COVIDOSE patients were as follows: Heart failure (I50·1, I50·2, I50·3, I50·4, I50·8, I50·9), history of percutaneous coronary intervention (Z98·61, Z95·5), history of coronary artery bypass graft (Z95·1), history of cerebrovascular accident with residual deficit (I69 and all subcategories), history of pulmonary hypertension (I27·0, I27·2), home oxygen dependence (Z99·81), interstitial lung disease (J84 and all subcategories), history of chronic obstructive pulmonary disease (J44·9), history of pneumonectomy (Z90·2), radiation therapy to lung (J70·1), HIV/AIDS (B20 and all subcategories), immunodeficiency (D81, D82, D83, D84), end-stage renal disease (N18·6), and obesity (E66·0, E66·1, E66·2, E66·8, E66·9). Cancers receiving active treatment were not interrogated for.

Using the International Classification of Diseases, 10^th^ Revision, Clinical Modification, Covid-19-relevant comorbidities were documented as follows: Heart failure (I50·1, I50·2, I50·3, I50·4, I50·8, I50·9), history of percutaneous coronary intervention (Z98·61, Z95·5), history of coronary artery bypass graft (Z95·1), history of cerebrovascular accident with residual deficit (I69 and all subcategories), history of pulmonary hypertension (I27·0, I27·2), home oxygen dependence (Z99·81), interstitial lung disease (J84 and all subcategories), history of chronic obstructive pulmonary disease (J44·9), HIV/AIDS (B20 and all subcategories), immunodeficiency (D81, D82, D83, D84), end-stage renal disease (N18·6), obesity (E66·0, E66·1, E66·2, E66·8, E66·9), diabetes mellitus (E10 and E11 and all subcategories), and hypertension (I10 and I11 and all subcategories). Cancers receiving active treatment were not interrogated for.

### SARS-CoV-2 Spike and RBD ELISA details

The SARS-CoV-2 Spike and Receptor Binding Domain (RBD) protein expression constructs were obtained courtesy of Florian Krammer and Patrick Wilson,^3^ and used in an enzyme-linked immunosorbent assay (ELISA) adapted from established protocols.^3^ Recombinant proteins were produced using a Chinese hamster ovary (CHO) cell line expression system and purified using metal-chelate affinity chromatography. Protein integrity was confirmed via SDS-PAGE gel.

Overnight, 96-well ELISA plates (Nunc MaxiSorp high protein-binding capacity plate; ThermoFisher) were coated at 4°C with 2 μg/mL of Spike or RBD protein suspended in Phosphate Buffered Saline (PBS) pH 7·4. Plates were blocked with 3% milk powder in PBS containing 0·1% Tween-20 for 1 hour at room temperature. Plasma and serum samples were heated at 56°C for 30 minutes to inactivate virus. Serial 1:3 dilutions of the samples were prepared in 1% milk in 0·1% PBS-Tween 20, and incubated in duplicate with the blocked plate for 2 hours at room temperature. After 3 washes in 0·05% PBS-Tween 20, a goat anti-human immunoglobulin (Ig)-Horseradish peroxidase (HRP) conjugated secondary antibody (SouthernBiotech, Catalog #2010-05, Lot #C2117-T819E), reactive to heavy and light chains of human IgG, IgM, and IgA, was diluted in 1% milk in 0·1% PBS-Tween-20, and added at 1:8000 for 1 hour at room temperature. Plates were washed 3 times with 0·1% PBS-Tween 20 before being developed with 3, 3’, 5, 5’-tetramethylbenzidine (TMB) substrate kit (ThermoFisher) at room temperature. The reaction was stopped after 15 minutes with 2M sulfuric acid. The optical density (OD) was read at 450 nm using a Synergy H4 plate reader (BioTek). The OD values for each sample were background subtracted. A positive control standard was prepared from plasma samples pooled from 6 Covid-19-infected patients, while plasma from an uninfected individual was used as a negative control standard. To account for variability between plates, the OD values were divided by the OD from the negative control from each plate, run at a 1:50 dilution. To quantify the amount of anti-Spike Ig and anti-RBD Ig in the sample, end-point titers were calculated as the linear interpolation of the inverse dilution at which the normalized OD value crossed a threshold of 1, representing the maximum OD measured for the negative control.

## Supplementary Results

### Re-dosing of Low-Dose Tocilizumab

In the originally developed re-dosing schema, re-dosing was guided strictly by CRP response. Six (18·8%) patients (2 in group A and 4 in group B) were re-dosed low dose tocilizumab. Five of 6 (83·3%) patients had continued clinical improvement demonstrated by decreasing requirement for supplemental oxygen and improving temperature curve. The remaining patient did not have a clinical response to two doses of low-dose tocilizumab. Three of 6 (50·0%) achieved the biochemical outcome of ≥25% reduction in CRP in the 24-48 hours following the first tocilizumab dose. In the final version of the protocol, re-dosing was guided by biochemical and clinical parameters (as defined by decreased supplemental oxygen requirement and improving fever curve determined by maximum temperature). No patients required re-dosing of tocilizumab under these conditions.

## Supplementary Figures and Table

**Figure S1. The COVIDOSE algorithm for inpatient administration of low-dose tocilizumab to patients with moderate or severe Covid-19**. Eligible COVIDOSE patients receive IL-6 axis-suppressing therapy. Patients have baseline clinical and laboratory-based risk stratification, and then receive an initial dose of tocilizumab. Usual clinical care continues for 24 hours after the initial tocilizumab dose, at which point repeat biochemical assessment is performed. If evidence of clinical decline and evidence of insufficient IL-6 axis suppression, COVIDOSE patients receive an additional dose of tocilizumab. Covid-19, novel coronavirus disease 2019; FiO_2_, fraction of inspired O_2_; IL-6, interleukin 6.

**Figure S2. Derivation of the retrospective control population from de-identified clinical data warehouse**. A retrospective control population of 41 patients (representing approximately 1·4% of positive tests and in line with the COVIDOSE population) was used for comparative analysis. SARS-CoV-2, severe acute respiratory syndrome coronavirus-2; PCR, polymerase chain reaction; IL-6, interleukin 6; mAb, monoclonal antibody; CRP, C-reactive protein; Covid-19, novel coronavirus disease 2019.

**Figure S3. Biobanked patients included in antibody analysis**. Patients were considered eligible if they had an infiltrate on their chest radiograph, a temperature ≥ 38°C, and CRP ≥ 40 mg/L within 24 hours. Patients were excluded if they had bacteremia or had concern for a serious bacterial infection, if they did not have an infiltrate on their chest radiograph, or if they did not provide blood samples available for analysis.

**Table S1. Demographic, clinical, and laboratory characteristics of the patients and controls analyzed for antibody responses**.

**Figure S4. Tocilizumab dose or remdesivir do not appear to alter antibody production**. Data shown in Figure 3 were sub-grouped by tocilizumab dose (blue: 40 mg, 80 mg, 120 mg, or 200 mg treated on COVIDOSE trial (n=19); red: 400 mg treated off-trial (n=4)) and controls were sub-grouped by remdesivir treatment (black: no remdesivir (n=11); gray: received remdesivir (n=26).

